# Systematic Review and Meta-analysis of Rate of Major Amputation Following Endovascular Intervention in Chronic Limb-Threatening Ischaemia

**DOI:** 10.1101/2022.12.20.22283746

**Authors:** Henry I Bergman, Hussein Elghazaly, Ankur Thapar, Alun H. Davies

**Affiliations:** Department of Surgery and Cancer, Imperial College London; Department of Surgery and Cancer, Imperial College London, London UK; Centre for Circulatory Health, Medical Technology Research Centre, Anglia Ruskin University, Chelmsford UK

## Abstract

**Background:** Despite significant improvements in endovascular technology and anaesthetic practice over the last 20 years, patients with CLTI remain at high risk of major limb amputation and overall mortality. The aim of this systematic review was to provide a contemporary review of the rate of major amputation and key clinical outcomes following all endovascular interventions in CLTI.

**Methods:** A systematic review and meta-analysis of prospective studies from 2010-2020 reporting the risk of major lower limb amputation in patients with CLTI. MEDLINE, EMBASE, the Cochrane Database for Systematic Reviews and ClinicalTrials.gov were searched for relevant studies by 2 reviewers. The primary endpoint was rate of major limb amputation at 1 year and 2 years. Meta-analysis of proportions was employed using the random effects model. Studies were quality assessed using the ROBINS-I tool. To investigate factors associated with major limb amputations, subgroup analyses and meta-regression for clinical-demographic and lesion characteristics were employed.

**Results:** A total of 28 studies, from 24 manuscripts were eligible for inclusion. These included a total of 49,756 patients. At one-year post-revascularisation, the pooled rate of major lower limb amputations at 1 year was 8.6% (95% CI 6.7% - 11.0%). At two years, the rate of major amputations was 11.1% (95% CI 7.6% - 16.0%). Subgroup analysis showed that in studies that mandated stent deployment for all patients, there was a significantly lower rate of major amputations of 5.1% (95% CI 4.7% - 5.5%). Meta-regression showed that none of the clinical-demographic and lesion characteristics were associated with major lower limb amputations, and no volume-outcome relationship was observed.

**Conclusions:** This review provides important benchmarking information on the outcomes of endovascular intervention in a frail CLTI cohort. This provides a realistic evaluation of risk to facilitate full informed consent and the setting of realistic expectations regarding the need for reintervention, major and minor amputation and overall mortality.

## Introduction

Critical limb-threatening ischaemia (CLTI) is the end stage of peripheral vascular disease, characterized by rest pain and/or tissue loss (Rutherford classification 4, 5 and 6)^1^. The incidence of peripheral arterial disease (PAD), and therefore CLTI, are increasing within the UK – with an aging and increasingly diabetic population being large drivers of disease prevalence^2,3,4^. These patients are well documented to have high peri-operative mortality, significant co-morbidity and disability^1^. Broadly, first line treatment modalities include open revascularization (bypass or endarterectomy), endovascular revascularization (angioplasty, stenting, atherectomy or lithotripsy) or a hybrid procedure^5^. Despite significant improvements in endovascular technology and anaesthetic practice over the last 20 years^5^, patients with CLTI are perceived to be at high risk of major limb amputation and overall mortality^1^.

In the UK, no NICE guidance exists for follow up after lower limb endovascular interventions ^6, 7^. The European Society of Vascular Surgery (ESVS) and European society of Cardiology (ESC) proposes a formal Duplex Ultrasound surveillance program with the aim of detecting clinically significant re-stenoses and facilitating reintervention prior to occlusion^8^. There is currently limited evidence in the literature from two non-randomized retrospective cohort studies to support implementing this program within the UK, which appear to demonstrate a reduction in major amputation rate following formal duplex surveillance^6,7^.

Regional clinical practice, utilization different endovascular technologies, surveillance strategies and patient selection for endovascular intervention vary significantly within the literature^5^. When considering these factors and the significant changes to practice over the last decade; to the best of the authors knowledge there is no clear consensus in the literature, or no contemporary review to delineate key outcomes following endovascular intervention. The aim of this systematic review was to provide a contemporary review of the rate of major amputation and key clinical outcomes following all endovascular interventions in CLTI.

## Methods

### Search Strategy

A comprehensive systematic literature search was conducted as per the Preferred Reporting Items for Systematic Reviews and Meta-Analyses (PRISMA) guidelines by two independent reviewers (HB and HE). Literature on the outcomes of patients with CLTI following any endovascular procedure from January 2010 to January 2020 were searched for in Medline, Embase, Cochrane database and ClinicalTrials.gov. The search terms included the key words “Endo” or “Angio” or “Stent” or “Atherectomy” or “Lithotripsy” and “Outcome” or “Amputation” or “Limb salvage”. As per the PRISMA guidelines additional studies were identified from the references of eligible literature.

### Eligibility criteria

Two independent authors (H.B and H.E) screened all literature identified from the search, conflicts were resolved by a third experienced author (A.T). Inclusion criteria included all prospective studies, with procedures performed from January 2010 onwards, on greater than 100 CLTI patients (Defined as rest pain or tissue loss), with a minimum of 1-year follow-up data available for major amputation. Exclusion criteria included studies performed outside of the 2010-2020 window, retrospective study methodology, hybrid procedures (Open and endovascular intervention) and procedures performed for intermittent claudication or aneurysmal disease. Case reports, review articles, articles conducted during the COVID-19 pandemic and invited commentaries were also excluded.

### Data extraction

Two independent authors (H.B and H.E) performed data extraction following sequential screening of titles, abstracts, and full texts. Two papers were unable to be retrieved despite contacting the authors and were therefore excluded. Baseline study, cohort and co-morbidity characteristics were recorded – these are summarized in **Table 2**.

**Table 1:** Characteristics of Included Studies.

| Study | Number of Patients | Number of Limbs | Number of Centres | Year of Publication | Type of Endovascular Treatment | Target Lesion Anatomy | Follow-Up |
| --- | --- | --- | --- | --- | --- | --- | --- |
| Abualhin 2019<sup>9</sup> | 211 | 211 | NS | 2019 | POBA, DCB | Femoral, BTK | 22 months |
| Ahn 2018 (POBA Cohort)<sup>10</sup> | 289 | 111 | 1 | 2018 | POBA | NS | 12 months |
| Ahn 2018 (Stent Cohort)<sup>10</sup> | 111 | 289 | 1 | 2018 | BMS, DES | NS | 24 months |
| Biasi 2016<sup>11</sup> | 201 | 201 | 1 | 2016 | NS | NS | 16 months |
| Bisdas 2016<sup>12</sup> | 642 | NS | 27 | 2016 | NS | NS | 12 months |
| Bondarenko 2014<sup>13</sup> | 164 | 193 | 1 | 2014 | NS | NS | 12 months |
| Brodmann 2020<sup>14</sup> | 328 | 422 | 45 | 2020 | POBA, DCB, Atherectomy | Femoral, BTK | 24 months |
| Desai 2020<sup>15</sup> | 186 | 186 | 1 | 2020 | POBA, DES, Atherectomy | BTK Only | 12 months |
| Elbadawy 2018<sup>16</sup> | 117 | 117 | 1 | 2018 | POBA | BTK Only | 12 months |
| Garcia 2017<sup>17</sup> | 201 | 201 | 47 | 2017 | POBA, Atherectomy | Femoral, BTK | 24 months |
| Geraghty 2020<sup>18</sup> | 180 | 180 | 1 | 2020 | POBA, BMS | Iliofemoral, BTK | 36 months |
| Giannopoulos 2020<sup>19</sup> | 1204 | NS | 52 | 2019-2021 | POBA, BMS, DCB, DES, Atherectomy | Femoral, BTK | 36 months |
| Giaquinta 2017<sup>20</sup> | 122 | 122 | 1 | 2017 | POBA, DES | BTK Only | 36 months |
| Hicks 2019<sup>21</sup> | 142 | 142 | 1 | 2019 | POBA, DCB, DES, Atherectomy | Femoral, BTK | 48 months |
| Iida 2017<sup>22</sup> | 351 | 351 | 23 | 2017 | BMS, DES | Femoral, BTK | 36 months |
| Khan 2020 (ACEi Cohort)<sup>23</sup> | 5558 | 5558 | NS | 2020 | POBA, BMS, Atherectomy | Iliofemoral, BTK | 60 months |
| Khan 2020 (Non-ACEi Cohort)<sup>23</sup> | 5773 | 5773 | NS | 2020 | POBA, BMS | Iliofemoral, BTK | 60 months |
| Lee 2020<sup>24</sup> | 172 | 172 | 1 | 2020 | POBA, BMS, DCB | NS | 24 months |
| Mustapha 2017<sup>25</sup> | 328 | 328 | 3 | 2017 | POBA, BMS, Atherectomy | Femoral, BTK | 12 months |
| Mustapha 2019 (Atherectomy Cohort)<sup>26</sup> | 4422 | 11295 | NS | 2019 | BMS | NS | 48 months |
| Mustapha 2019 (Stent Cohort)<sup>26</sup> | 11295 | 4422 | NS | 2019 | Atherectomy | NS | 48 months |
| Mustapha 2019 (PTA Cohort)<sup>26</sup> | 10904 | 10904 | NS | 2019 | POBA | NS | 48 months |
| Perlander 2020<sup>27</sup> | 117 | 117 | 3 | 2020 | POBA, BMS | Femoral, BTK | 24 months |
| Reijnen 2019<sup>28</sup> | 156 | 194 | NS | 2019 | DCB | Femoral, BTK | 12 months |
| Riambau 2020<sup>29</sup> | 148 | 148 | 10 | 2019 | DCB | Femoral, BTK | 12 months |
| Siracuse 2016<sup>30</sup> | 5387 | 5387 | NS | 2016 | POBA, BMS, Atherectomy | Femoral | 60 months |
| Teichgraber 2019<sup>31</sup> | 164 | NS | 9 | 2019 | DCB | BTK Only | 12 months |
| Vierthaler 2015<sup>32</sup> | 883 | 883 | 23 | 2013 | POBA, BMS, DCB, DES, Atherectomy | Iliofemoral, BTK | 12 months |

**Key: POBA = Plain Old Balloon Angioplasty; DCB = Drug Coated Balloon; BMS = Bare Metal Stent; DES = Drug Eluting Stent; BTK = Below the Knee, NS=not stated.**

**Table 2:** Summary of Baseline Lesion Characteristics and Patient Demographics in Included Studies.

| Study | Lesion Length (mm) | Percentage of CTO (%) | Age (Years) | Smokers (%) | Female (%) | Diabetes (%) | Hypertension (%) | Dyslipidaemia (%) | IHD (%) | ESRD (%) |
| --- | --- | --- | --- | --- | --- | --- | --- | --- | --- | --- |
| Abualhin 2019<sup>9</sup> | NS | NS | 73 | NS | 28% | 33% | NS | NS | 48% | NS |
| Ahn 2018 (POBA Cohort)<sup>10</sup> | NS | NS | NS | NS | NS | NS | NS | NS | NS | NS |
| Ahn 2018 (Stent Cohort)<sup>10</sup> | NS | NS | NS | NS | NS | NS | NS | NS | NS | NS |
| Biasi 2016<sup>11</sup> | NS | NS | 73 | NS | 30% | 64% | NS | NS | NS | NS |
| Bisdas 2016<sup>12</sup> | NS | NS | 75 | NS | 37% | 48% | NS | NS | 46% | 10% |
| Bondarenko 2014<sup>13</sup> | NS | NS | 64 | NS | 54% | 100% | NS | NS | 15% | NS |
| Brodmann 2020<sup>14</sup> | 82 | 27% | 71 | 57% | 39% | 61% | 87% | 61% | 44% | 9% |
| Desai 2020<sup>15</sup> | NS | NS | 72 | 62% | 43% | 66% | 75% | 62% | 60% | 16% |
| Elbadawy 2018<sup>16</sup> | NS | NS | 67 | 76% | 35% | 90% | 20% | 39% | 26% | 12% |
| Garcia 2017<sup>17</sup> | 72 | 30% | 72 | 36% | 50% | 69% | 92% | 76% | 32% | 23% |
| Geraghty 2020<sup>18</sup> | NS | 48% | 74 | 62% | 33% | 66% | NS | 78% | 56% | 25% |
| Giannopoulos 2020<sup>19</sup> | 127 | 43% | 61 | 64% | 37% | 71% | 93% | 84% | 62% | NS |
| Giaquinta 2017<sup>20</sup> | 53 | 68% | 71 | 66% | 36% | 53% | 80% | NS | 74% | 33% |
| Hicks 2019<sup>21</sup> | NS | NS | 65 | 57% | 38% | 100% | 91% | 71% | 47% | 15% |
| Iida 2017<sup>22</sup> | NS | NS | 74 | 15% | 34% | 74% | NS | NS | 40% | 55% |
| Khan 2020 (ACEi Cohort)<sup>23</sup> | NS | NS | NS | 73% | 43% | 67% | 94% | NS | 31% | 7% |
| Khan 2020 (Non-ACEi Cohort)<sup>23</sup> | NS | NS | NS | 72% | 44% | 57% | 84% | NS | 27% | 12% |
| Lee 2020<sup>24</sup> | NS | NS | 73 | 37% | 35% | 100% | 84% | 51% | 36% | 42% |
| Mustapha 2017<sup>25</sup> | 156 | 51% | 70 | 71% | 36% | 64% | 90% | 91% | 44% | 7% |
| Mustapha 2019 (Atherectomy Cohort)<sup>26</sup> | NS | NS | 74 | 21% | 48% | 61% | 80% | 30% | 55% | 34% |
| Mustapha 2019 (Stent Cohort)<sup>26</sup> | NS | NS | 74 | 25% | 50% | 52% | 74% | 27% | 48% | 31% |
| Mustapha 2019 (PTA Cohort)<sup>27</sup> | NS | NS | 74 | 16% | 48% | 63% | 73% | 27% | 52% | 40% |
| Perlander 2020<sup>27</sup> | NS | 65% | 75 | 18% | 46% | 36% | NS | NS | 32% | 9% |
| Reijnen 2019<sup>28</sup> | 139 | 41% | 72 | NS | 44% | 55% | 85% | 62% | 18% | 20% |
| Riambau 2020<sup>29</sup> | 77 | 54% | 73 | 54% | 32% | 72% | 87% | 57% | 39% | NS |
| Siracuse 2016<sup>30</sup> | NS | NS | 67 | 84% | 41% | 47% | 92% | NS | 36% | 5% |
| Teichgraber 2019<sup>31</sup> | 71 | 43% | 75 | 45% | 34% | 80% | 90% | 55% | 35% | NS |
| Vierthaler 2015<sup>32</sup> | NS | NS | NS | 74% | 42% | 61% | 90% | NS | 35% | 20% |

**Key: NS = Not Stated; CTO = Chronic Total Occlusion; IHD = Ischaemic Heart Disease; ESRD = End-Stage Renal Disease.**

### End points

The primary end point was the rate of major lower limb amputation (defined as an above ankle amputation). Secondary end points were primary, primary assisted and secondary patency, all-cause mortality, revascularization rate, Major adverse cardiovascular and cerebrovascular event (MACCE) rate, Quality of Life (Assessed by either EQ-5D, VascuQol-25 or VascuQol-6 questionnaires) and Minor amputation rate. For studies reporting outcomes as limb salvage these were converted into major amputation rates. All outcomes were assessed at 12 months and where possible also recorded at 24-months. All end points and sub-group analyses were pre-specified in the review protocol.

### Study quality

The methodology of eligible studies was evaluated using the ROBINS-I (Risk of Bias in Non-Randomized Studies of Interventions) tool to assess risk of bias in 7 key domains, and giving an overall score for quality (Critical, High, Moderate or Low risk of bias).

### Statistical analysis

Analysis, meta-analysis and meta-regression were performed using R Version 4.0.5 and RStudio 2022.07.1+554 (R Foundation for statistical computing – Vienna, Austria), using a random-effects model to compensate for heterogeneity between studies. Heterogeneity was measured with I^2^ values. The *“meta”* package was used to perform a meta-analysis of proportions for included studies. Pooled estimates were presented as percentage event rates for each pre-specified primary and secondary end point with 95% confidence intervals (CI).

Subgroup analysis was performed for below-knee interventions and primary stenting. In addition, where sub-group analysis was not possible, meta-regression was performed which to investigate any statistically significant effect of study characteristics, cohort demographics (specifically including number of patients treated, patient sex and proportion of patients with chronic total occlusion) or patient comorbidities on any of the specified end points.

## Results

### Database Search Results

Database searches identified a total of 4,065 results. A total of 28 studies, from 24 manuscripts were eligible for inclusion. These included a total of 49,756 patients. **Figure 1** outlines the PRISMA flowchart for study selection.

**Figure 1:**
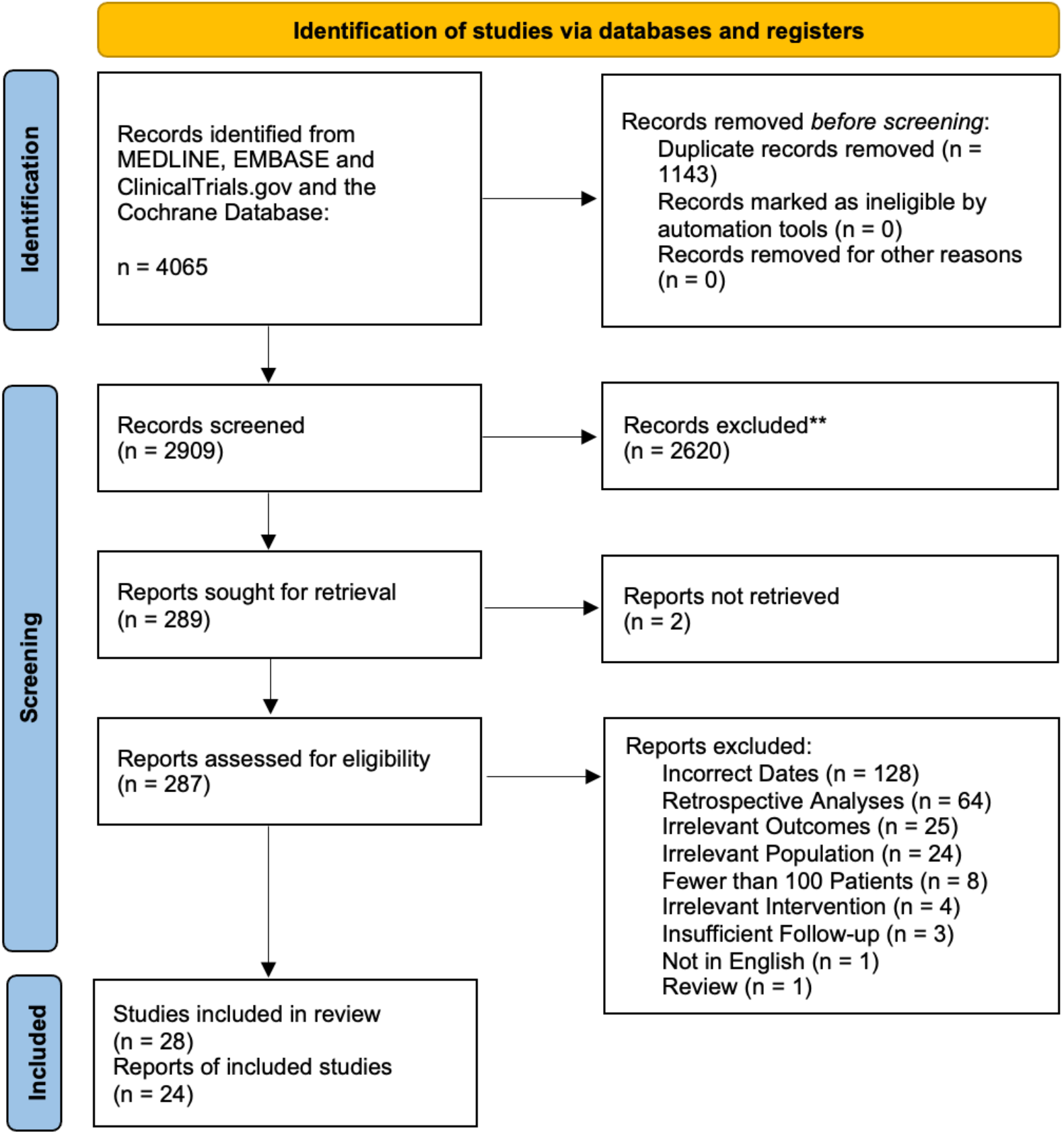
PRISMA Flowchart for Study Selection.

### Summary of Included Studies

**Table 1** summarises the characteristics for included studies. Briefly, all studies were prospective registries, prospective cohort studies or single arms of small randomised control trials (RCTs) were. The smallest study included 111 patients and the largest study followed up 11,295 patients. All studies followed patients up for at least 1 year; the longest follow-up period was 5 years, which occurred in 3 studies. Amongst included manuscripts, 10 were multicentre studies. All endovascular procedures were conducted between January 1^st^, 2010 and December 31^st^, 2019.

### Baseline Characteristics and Heterogeneity

A summary of baseline lesion and patient characteristics in included studies is illustrated in **Table 2**. As expected, due to the high-risk patient population, cardiometabolic risk factors were prevalent amongst participants in included studies. However, a high degree of clinical heterogeneity was also noteworthy. For instance, the anatomical location of the target lesion varied widely within the studies; 3 studies only included patients exclusively treated for infrapopliteal lesions (below the knee, BTK). Only four of the included studies included patients who received iliac artery interventions. In addition to anatomical differences, the type of endovascular procedure performed varied substantially between each study, and few studies reported outcomes for each device separately. Furthermore, although target lesion length was only reported in a minority of studies included in this review, there was a wide range (53mm – 152mm) in average target lesion length. Similarly, the proportion of chronic total occlusions (CTO, as opposed to stenoses) also showed noticeable heterogeneity (range 27% to 68%). Moreover, apart from hypertension, the prevalence of cardiovascular risk factors, ischaemic heart disease (IHD) and end-stage renal disease or dialysis (ESRD) was widely varied between the included studies.

### Risk of Bias

Overall, there was a moderate risk of bias in included studies according to the ROBINS-I tool. A risk of bias summary table is presented in **Figure 2**. Notably, apart from 2 manuscripts (Giaquinta et al. and Reijnen et al.), there was a high risk of selection and allocation bias in the included studies. Moreover, there was a moderate-to-high risk of bias due to confounding in 13 of the included studies. Since major lower limb amputation is an objective endpoint, it was deemed a low risk of bias in the ‘measurement of outcome’ domain for all included studies. Similarly, all included studies were deemed low risk of bias with regards to deviation from protocol, since endovascular procedures are short, and do not require patient adherence or compliance with therapy. No studies reported the compliance with best medical therapy, therefore the confounding effect of this is unknown. Owing to the nature of study designs and consequent low attrition rates in included studies, we deemed a low risk of bias due to ‘missing data’ in most studies.

**Figure 2:**
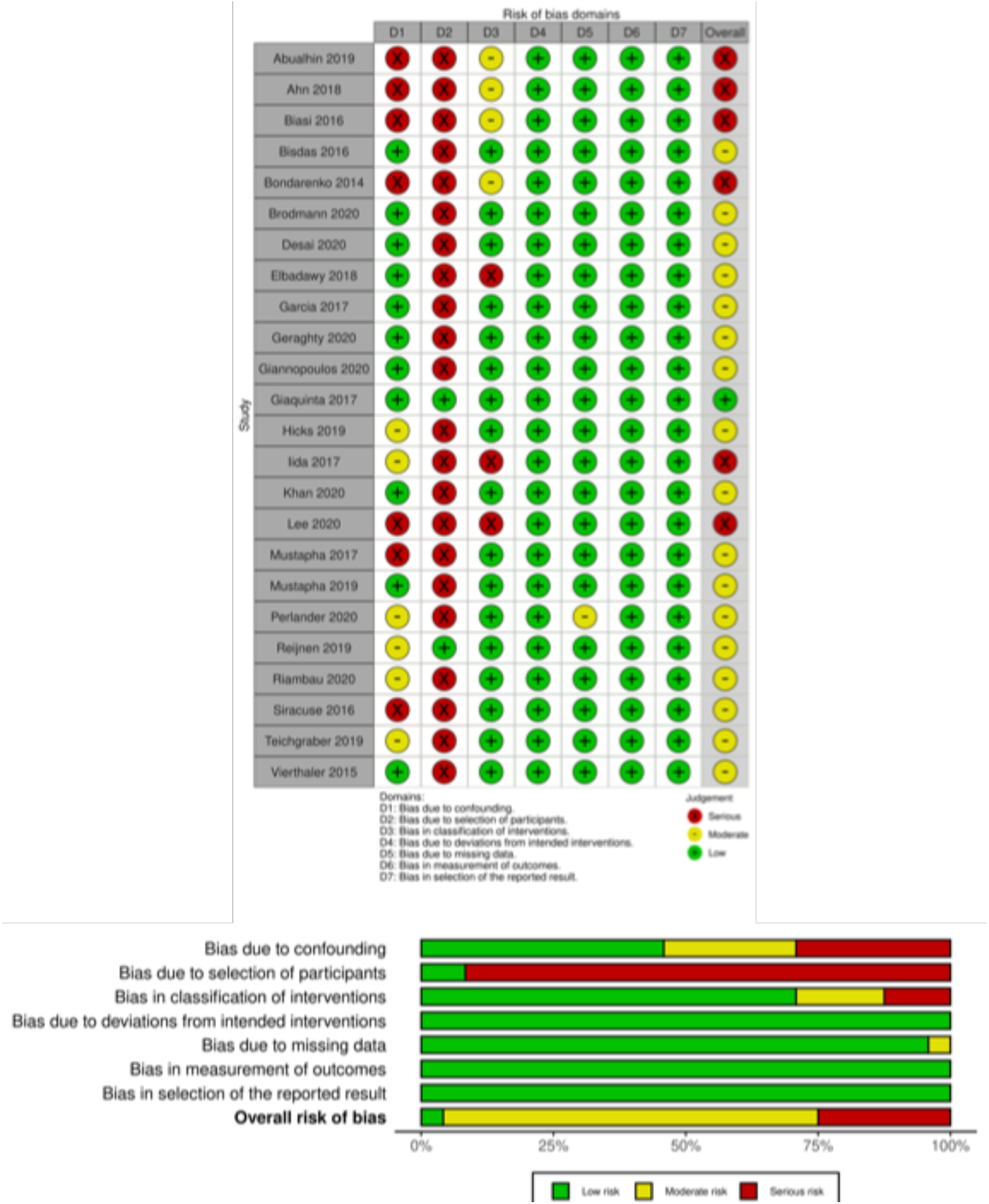
ROBINS-I. All bias domains were deemed as “Low Risk”, “Moderate Risk” or “Serious Risk” according to the ROBINS-I tool. Confounding bias refers to bias introduced due to imbalances of confounding variables including demographics, co-morbidities and lesion/ procedural characteristics. Bias due to participant selection denotes allocation bias that is introduced when assigning participants to their respective study arms. Bias due to classification of intervention encompasses the definition of interventions received by participants and whether intervention status label could have been affected by knowledge of the outcome. Deviation from intended intervention denotes nonconformity of interventions received by patients to the study’s original protocol. Missing data investigates bias of outcomes due to unbalanced follow-up in each study arm and imbalances in patients lost to follow-up. Measurement of outcome investigates whether x knowledge of the intervention received may have influenced the outcome. Selection of reported results investigates bias investigates whether authors have reported results in a manner that biases the findings of the study. This includes reporting multiple outcome measurements within the outcome domain, reporting multiple analyses of the intervention-outcome relationship, and reporting different subgroups.

### Meta-Analysis

A summary of the conducted meta-analyses is presented in **Table 3**.

**Table 3:** Summary of Meta-Analyses.

|  | 1 Year |  |  | 2 Years |  |  |
| --- | --- | --- | --- | --- | --- | --- |
|  | N | Effect Size Estimate (RE) | 95% CI | N | Effect Size Estimate (RE) | 95% CI |
| Major Lower Limb Amputations | 23 | 8.6% | 6.7% - 11.0% | 10 | 11.1% | 7.6% - 16.0% |
| Primary Patency | 13 | 77.3% | 71.1% - 82.5% | 5 | 53.5% | 32.9% - 73.0% |
| Primary Assisted Patency | 4 | 80.5% | 72.2% - 86.8% | 2 | 63.8% | 44.6% - 79.4% |
| Secondary Patency | 8 | 82.5% | 76.8% - 87.1% | 4 | 73.5% | 60.2% - 83.6% |
| All-Cause Mortality | 16 | 12.5% | 10.1% - 15.4% | 8 | 25.2% | 21.7% - 29.1% |
| Re-Intervention | 10 | 10.2% | 5.7% - 17.5% | 3 | 19.4% | 10.9% - 32.1% |
| Minor Amputation | 8 | 16.7% | 11.6% - 23.3% | 2 | 23.5% | 3.8% - 70.3% |

### Primary Outcome: Rate of Major Lower Limb Amputations

At one-year post-revascularisation, 23 studies with 37,888 patients reported the rate of major lower limb amputations. The pooled rate of major lower limb amputations at 1 year was 8.6% (95% CI 6.7% - 11.0%). There was significant statistical heterogeneity amongst included studies (I^2^ = 97%, *p* < 0.01). The Forest plot for this analysis is shown in **Figure 3**. At two years, 10 studies with 29,126 patients reported the rate of major lower limb amputations. The pooled rate of major lower limb amputations was 11.1% (95% CI 7.6% - 16.0%). There was significant statistical heterogeneity amongst included studies (I^2^ = 98%, *p* < 0.01). The Forest plot for this analysis is shown in **Figure 4**. There was no evidence of publication bias by visual inspection of the funnel plot or by Egger’s test **(Supplemental Material S1)**.

**Figure 3:**
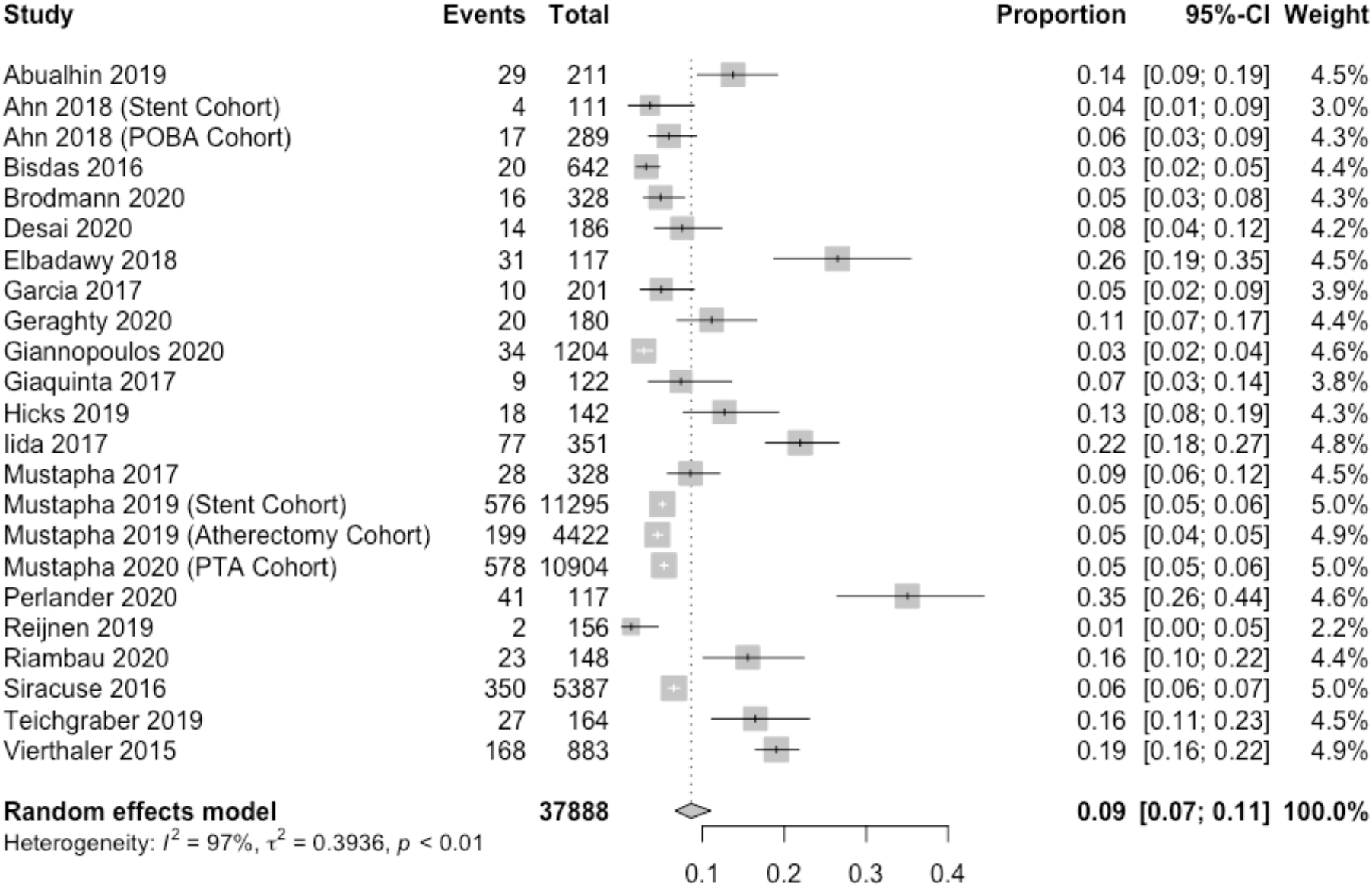
Forest Plot for Risk of Lower Limb Amputation at 1 Year.

**Figure 4:**
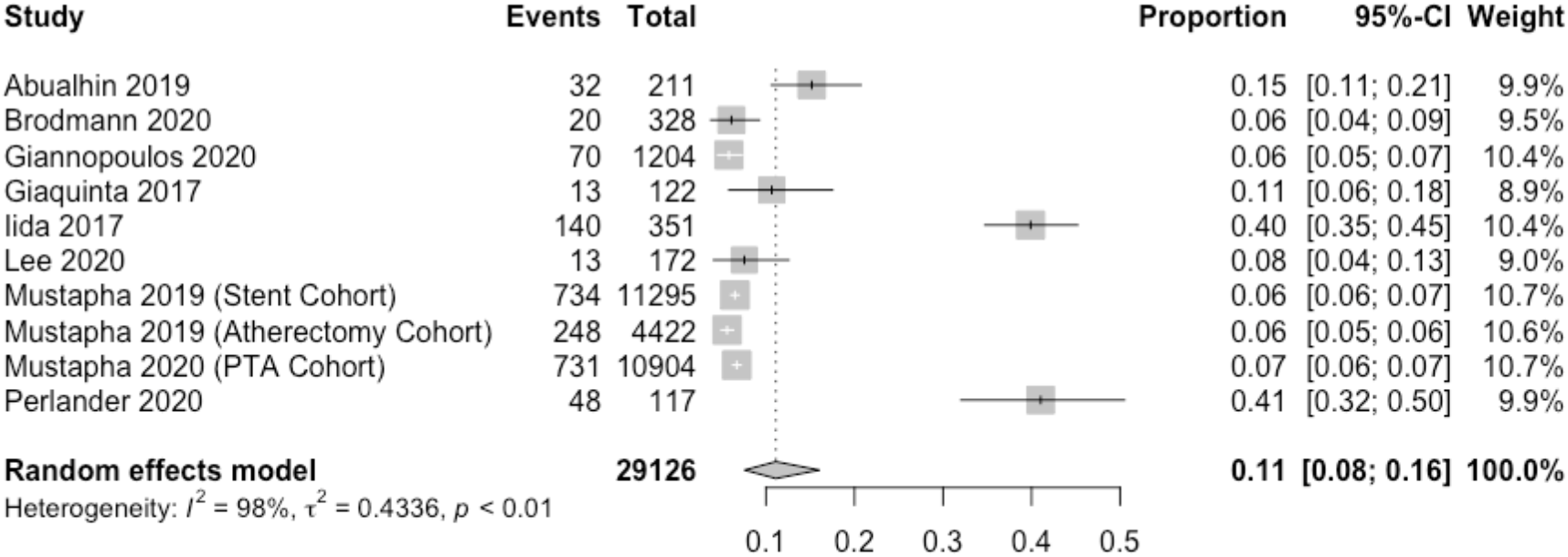
Forest Plot for Risk of Lower Limb Amputation at 2 Years.

### Subgroup Analysis

Three studies included 425 patients who were revascularised for exclusively infrapopliteal (BTK) lesions. A subgroup analysis of these patients showed that the pooled rate of major lower limb amputations at 1 year was 8.2% (95% CI 6.3% - 10.6%). There was significant statistical heterogeneity amongst included studies (I^2^ = 92%, *p* < 0.01). A Forest plot for this analysis is shown in **Figure 5**.

**Figure 5:**
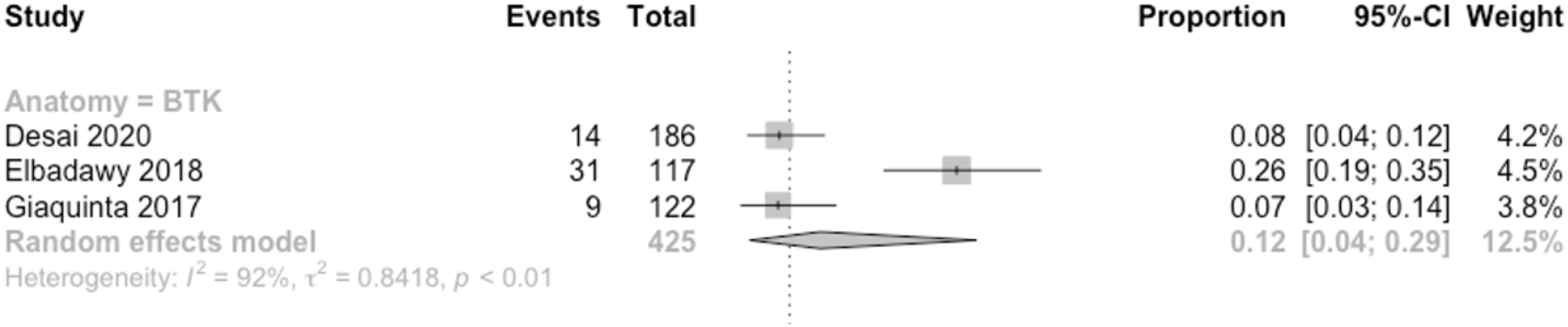
Forest plot for risk of lower limb amputation at 1 year in studies where all patients were revascularised for exclusively infrapopliteal (BTK) lesions.

Furthermore, three of the included studies mandated stent deployment for all patients. These studies included a total of 11,528 patients. The anatomy of the target lesion was variable within this cohort; one study included only infrapopliteal lesions and the other two contained a combination of iliac, femoral and BTK lesions. A subgroup analysis of these 3 studies showed that the pooled rate of major lower limb amputations at 1 year was 5.1% (95% CI 4.7% - 5.5%). No statistical heterogeneity was noted amongst included studies (I^2^ = 0%, *p* = 0.41). A Forest plot for this analysis is shown in **Figure 6**.

**Figure 6.**
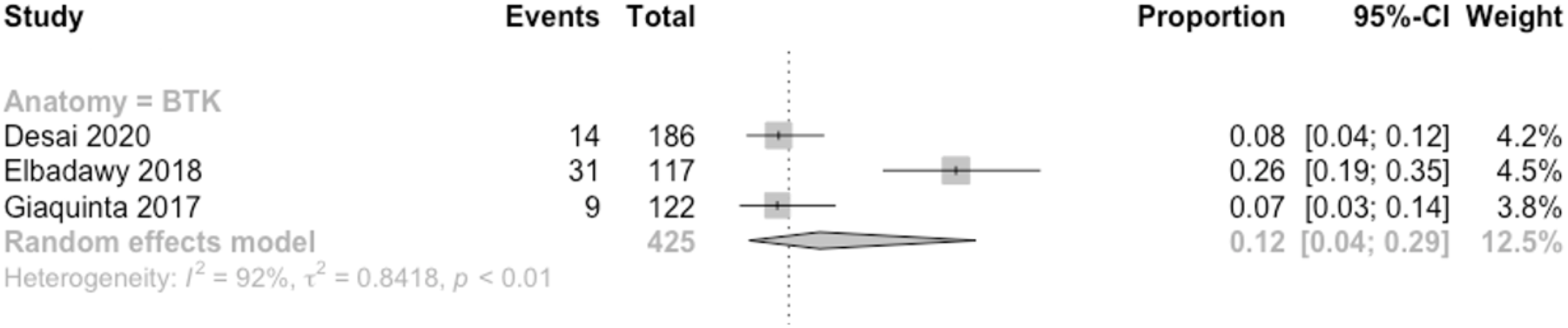
Forest plot for risk of lower limb amputation at 1 year in studies that mandated stent deployment for all patients.

### Meta Regression

Results of the meta-regression on factors influencing the rate of major lower limb amputation at one year is summarised in **Table 4**.

**Table 4:**
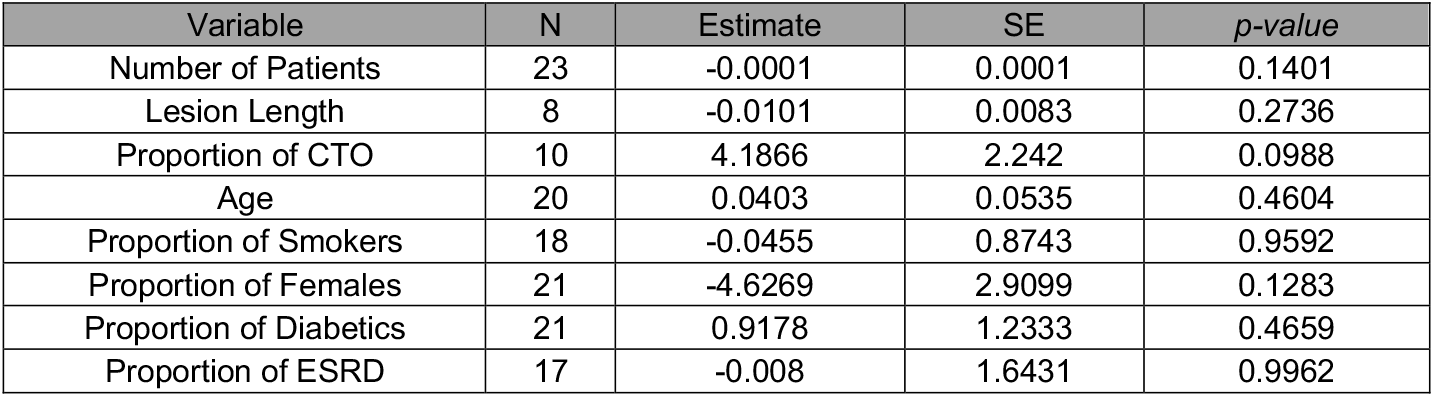
Results of Meta-Regression Analysis for Lower Major Limb Amputations at 1 year.

Notably, there was no significant correlation between the number of patients in a study and risk of major lower limb amputation at one year (Estimate -0.0001 ± 0.0001, *p* = 0.1401). A bubble plot of this analysis is shown in **Figure 7A**.

**Figure 7:**
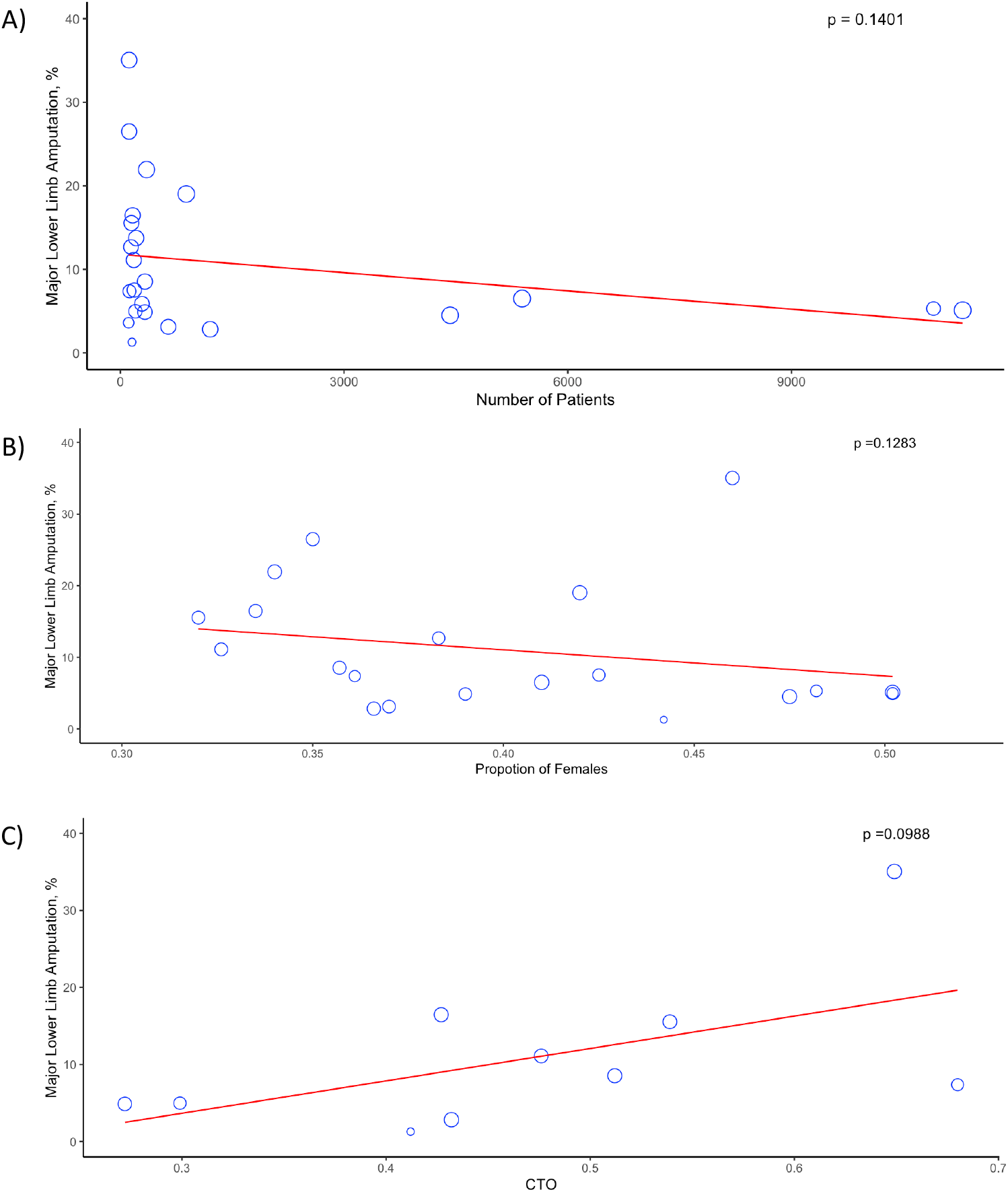
Bubble Plots for Meta-Regression Analyses for Major Lower Limb Amputation at 1 Year. A) Meta-regression for number of patients; B) Meta-regression for proportion of females; C) Meta-regression for proportion of patients with chronic total occlusion (CTO).

With regards to difference between males and females, no correlation was noted in the meta-regression of proportion of females and risk of major lower limb amputation at one year *Estimate -4.6269 ± 2.9099, *p* = 0.1283). A bubble plot of this analysis is shown in **Figure 7B**.

Similarly, there was no relationship between the proportion of patients with chronic total occlusions (CTO) and risk of major lower limb amputation at one year (Estimate 4.1866 ± 2.242, *p*= 0.0988). A bubble plot of this analysis is shown in **Figure 7C**.

None of the variables investigated in meta-regression showed a significant association with risk of major lower limb amputation at one year (**Table 4**).

### Secondary Endpoints

Amongst included studies, there was insufficient data to perform meta-analysis for MACCE rate or quality of life outcomes. Meta-analysis was performed for one- and two-year outcomes for all remaining secondary endpoints. **Table 3** summarises the outcomes of these analyses. Forest plots for the analyses are included in the **Supplementary Material (S2-S13)**.

## Discussion

This study established the contemporary rate of major amputation following endovascular intervention for CLTI to be 9% and 11% at 1- and 2-years follow-up respectively. This is an important figure which is relevant to providing informed consent and facilitates international benchmarking for endovascular practitioners. In addition, it found a similar risk in below the knee interventions, which were previously thought of as high risk, possibly reflecting an improvement in the treatments available for this subgroup. Interestingly, patients undergoing primary stenting showed a significant reduction in major amputation rate to 5% at 1 year. This figure should be regarded as hypothesis generating, as there may be other differences (e.g. antiplatelet therapy and surveillance protocols) that may have biased this result. Reintervention rates were established at 10% and 25% at 1- and 2-years respectively. This high rate of reintervention does suggest patients should be followed up for 2 years if fit enough for a further procedure. It should be recognized that 25% of patients are dead at 2 years, and that this extremely frail group of patients may not be candidates for intensive ultrasound surveillance, unlike younger fitter patients. Finally, no volume-outcome relationship was observed, in contrast to aortic aneurysm^33^ and carotid surgery^34^ - to our knowledge this has not been previously observed in the literature.

Historical metanalyses quote rates of up to 14% for 1-year amputation rate following endovascular intervention^35^, indicating a possible change in superior technology, better medical therapy, ore more intensive surveillance when compared with the contemporary outcomes elicited in this review. Further research to establish the temporal trends of these outcomes over the last decade would be needed to determine whether rates of amputation have changed significantly.

UK National Vascular Registry data reports 30-day post-interventional amputation rate as 1.2% and 9.5% for elective and emergency endovascular revascularization respectively (combined weighted average of 3.5%)^36^. Indeed, 30-day outcomes from the American NSQUIP registry are similar – reported as 4.1%^37^. Despite being very large data sets, the limited case ascertainment (Only 46% for NVR) make these difficult to compare to real world practice or to extrapolate to this review’s reported 1 and 2 year outcomes. These registries do demonstrate that at least half of limbs lost, occur in the first month after the procedure.

The strengths of this study were that included patients actually treated between January 2010 and January 2020 and therefore reflects recent international outcomes. It excluded patients managed during the COVID-19 pandemic, during which time the practice of many vascular units temporarily changed. The predominant study type included in the review were prospective registries, with few small randomized trials, therefore there was excellent classification, measurement and reporting of outcomes, with very little missing data at follow-up intervals.

The limitations of this study were that it could not perform individual patient data analyses on all informative subgroups. Instead exploratory meta-regression analyses were performed on several clinically relevant subgroups, e.g. gender, dialysis and chronic total occlusion. Of these only the last, showed a trend towards significance. It was not possible to extract atherectomy as an additional sub-group, which would have been informative. With the exception of two^20,28^, all included studies suffered from selection bias, a limitation of the available data. It is likely that those that publish data, achieve better outcomes, and hence the figures quoted represent what should be achievable, rather than what every practitioner is achieving. However, the higher 95% confidence intervals for amputation outcomes (11% at 1 year and 16% at 2 years) should provide a useful benchmark for practitioners and regulators in this field. Finally, there was significant clinical, methodological and statistical heterogeneity between the studies, consistent with real world practice.

This review provides important benchmarking information on the outcomes of endovascular intervention in a frail CLTI cohort. Crucially this provides a realistic evaluation of risk to facilitate full informed consent and the setting of realistic expectations regarding the need for reintervention, major and minor amputation and overall mortality. On the healthcare provision and governmental level this review provides important prognostic information regarding the rate of conversion of CLTI patients to major amputees. Evidence for duplex surveillance after endovascular therapy is limited to 2 comparative studies^6,7^, therefore, another research priority should be the cost-effectiveness of routine duplex surveillance versus clinical surveillance after intervention.

Further research in this field would be valuable to patient centered outcomes following all lower limb interventions (open, hybrid, endovascular, primary amputation and conservative). In addition to conventionally reported endpoints (major amputation rate, patency and reintervention), patient reported outcomes such as Quality of life (QoL), illness perception, formal and informal carer requirement, and Barthel index of independence of activities of daily living, would be valuable to understand the wider impact of those undergoing surgical therapy for peripheral arterial disease.

## Data Availability

All data produced in the present study are available upon reasonable request to the authors

## Acknowledgements

**N/A**

## Sources of Funding

**None**

## Disclosures

**None**

## Notes

### Competing Interest Statement

The authors have declared no competing interest.

### Funding Statement

This study did not receive any funding

